# Monocytic Subsets and Their Signature Genes Differentially Impact Cortex and Cognition in First-Episode Schizophrenia

**DOI:** 10.1101/2021.09.13.21262813

**Authors:** Song Chen, Keerthana Chithanathan, Fengmei Fan, Meihong Xiu, Hongzhen Fan, Yimin Cui, Ping Zhang, Ting Yu, Fude Yang, Baopeng Tian, L. Elliot Hong, Yunlong Tan, Li Tian

## Abstract

Accumulating evidence supports involvement of innate immunity in the pathophysiology of schizophrenia. Monocytes are a highly heterogeneous population, subcategorized into classical (CD14++CD16−), intermediate (CD14++CD16+) and nonclassical subsets (CD14+CD16++). How monocytic subsets may shape brain structures and functions remains unclear. The primary goal of this cross-sectional study was to investigate the inter-relationships among monocytic subsets and their specific transcriptomic profiles, cerebral cortical thickness, and cognitive functions in first-episode schizophrenia (FES) patients. We performed whole-blood RNA sequencing (RNAseq) in 128 FES patients and 111 healthy controls (HCs) along with MATRICS Consensus Cognitive Battery (MCCB) measurement, as well as neuroimaging and flow cytometry among partial participants. RNAseq revealed significantly changed expressions of 54 monocytic signature genes in FES patients compared to HCs, especially for intermediate and nonclassical monocytic subsets, with the most outstanding alterations being downregulated S100 Calcium Binding Protein A (*S100A*) and upregulated Interferon Induced Transmembrane Protein (*IFITM*) family members, respectively. The percentage of nonclassical monocytes was decreased in FES patients. Cortical thicknesses and MCCB performance were expectantly reduced in FES patients too. Interestingly, negative inter-relationships of monocytic signature genes with both cortical thicknesses and cognition were found in HCs, which were weakened or even reversed in FES patients. Furthermore, the lateral occipital cortex fully mediated the negative effect of a classical monocytic gene Ribonuclease A Family Member 2 (*RNASE2*) on visual learning in patient group. This study suggests that monocytic dysfunctions play an essential role in cognitive deficit of schizophrenia, and their subtypes should be considered in future research.

**One Sentence Summary:** Dysfunctions of monocytic subsets play an essential role in cortex and cognitive deficit of schizophrenia

## INTRODUCTION

The immune system has been found to interact with the central nervous system (CNS) in various cognitive and behavioral functions, the dysregulation of which contributes to chronic low-grade inflammation in mental disorders, such as schizophrenia, bipolar disorder, depression, or Alzheimer’s disease (*1, 2*). We recently observed that allostatic load, a composite index (including immune factors) of stress maladaptation, negatively contributed to cortical and cognitive deficits (*3*) and was associated with enlargement of the choroid plexus – an important cerebral portal for circulatory immune cells – in treatment-naïve first-episode schizophrenia (FES) patients (*4*).

Innate immune responses provide the first line of defense against invading pathogens and activate adaptive host defense mechanisms. Microglia and monocytes/macrophages are the primary innate immune phagocytes in the CNS parenchyma and peripheral tissues, respectively; monocyte-derived macrophages also reside at borderlines between the CNS and periphery, such as the meninges, perivascular spaces, and choroid plexuses (*5, 6*). Microglia share many myeloid features with monocytes and monocyte-derived macrophages, which all have been implicated in the pathophysiology of schizophrenia (*7, 8*). For instance, monocytes and macrophages can infiltrate into the stroma of choroid plexus and brain tissue in some pathological contexts and augment glia-mediated neuroinflammation, resulting in impaired synaptic plasticity and psychotic symptoms (*9–12*).

Monocytes are notably a heterogeneous population, which is reflected for example by the relative expression levels of CD14 and the low-affinity Fcγ-Ⅲ receptor (CD16) that namely divide monocytes into the classical (CD14++CD16−), intermediate (CD14++CD16+) and nonclassical subsets (CD14+CD16++) (*13, 14*). CD14++CD16− classical monocytes account for ~85% of peripheral blood monocytes, whereas CD14++CD16+ intermediate and CD14+CD16++ nonclassical subsets make up only ~5% and ~10%, respectively (*13, 15*). Each of these subsets is regarded as phenotypically distinct, carrying different signature genes and playing diverse roles in a myriad of chronic inflammatory and autoimmune conditions, including but not limited to Alzheimer’s disease and multiple sclerosis (*16–19*). CD14++CD16− classical monocytes have the highest phagocytic capacity and a predominant antimicrobial function (*13, 20*). By contrast, CD14+CD16++ nonclassical monocytes have been generally viewed as anti-inflammatory and patrol along the vasculature thereby playing a critical role in maintaining vascular homeostasis (*18*). CD14++CD16+ intermediate monocytes presumably represent a transitional stage between the classical and nonclassical monocytes, sharing some phenotypic and functional characteristics of both subsets (*15*), and are thought to be mainly involved in antigen processing and presentation to induce T-cell proliferation (*13, 14, 21*). They also have significant inflammatory capacity and increase oxidative stress (*22*).

The relationship between schizophrenia and monocytes has attracted increasing interest in recent years. For instance, a higher monocyte count was observed in patients with schizophrenia and related disorders (*23*); increased sizes or numbers of monocytic subcellular organelles, including the nucleolus, mitochondria and lysosomes, were observed in schizophrenic patients as compared to normal controls (*24*). Monocytic transcriptomes of patients with schizophrenia or bipolar disorder had stronger inflammatory gene expression fingerprints (*25*). Studies including ours have also found altered levels of toll-like receptors (TLR) and proinflammatory cytokine responses such as IL-1β in monocytes following the stimulation of monocytic TLR4/TLR5 and ErbB in schizophrenia (*24, 26, 27*). Another study revealed that monocytic TLR-4 expression was negatively associated with brain volumes of frontal and anterior cingulate regions in FES patients (*28*). Cortical thickness reduction, which reflects neuronal/glial dystrophy and/or loss of synapses, is a consistent structural MRI finding in schizophrenia by others and us (*3, 29, 30*). However, no study has investigated monocytic subsets so far and nothing is known whether and how monocytic subsets may shape brain structures and functions in both normal and schizophrenic conditions. Thereby, the primary aim of this study was to investigate the inter-relationships among monocytic subsets and their specific transcriptomic profiles, cortical thickness, and cognitive functions in FES patients.

## MATERIALS AND METHODS

### Ethical approval and participants

The present study used a cross-sectional research design. This study complied with the Declaration of Helsinki with regard to an investigation in humans, and the study protocol was approved by the Medical Ethical Committee of Beijing Huilongguan Hospital. All of the participants gave written informed consent before the initiation of study procedures.

FES patients (*n* = 128) were enrolled in Beijing Huilongguan Hospital from 2016 to 2018, and the inclusion criteria were: 1) diagnosis of schizophrenia based on the Structured Clinical Interview for DSM-IV (SCID), which was administered by two experienced psychiatrists; 2) Han nationality and aged 18-55 years old; 3) illness duration ≤36 months; 4) education equal or greater than 8 years; 5) right handedness, and physically healthy in the past; 6) cumulative exposure to psychotropic drugs ≤ 14 days; 7) receiving no immunomodulators, immune-suppressive or anti-inflammatory agents in the past 6 months; 8) no substance and alcohol abuse/dependence.

Healthy controls (HCs) (*n* = 111) were recruited from the local community also from 2016 to 2018, and assessed for personal history of mental disorders according to the standardized SCID diagnostic assessment. Individuals were excluded if they had first-degree relatives with a diagnosis of psychosis. The other general criteria were the same to FES patients.

### Blood sample collection, RNA sequencing (RNAseq) and data analysis

Whole blood (5 ml) was collected between 7 and 9 AM after overnight fasting using PAXgeneTM blood RNA tubes (Applied Biosystems, USA). Tubes were shaken vigorously for at least 10 seconds after sampling and immediately stored at −80°C. Total RNAs were extracted using Mag-MAX^TM^ RNA Isolation Kit (Applied Biosystems, USA) by following the manufacturers’ instructions. RNAs were quantified and assessed for purity by optical density ratios of 260nm/280nm and 260nm/230nm using NanoDrop spectrophotometry, and samples (1 µg) were immediately shipped on dry ice to the laboratory of the Beijing Genomics Institute (BGI), China for sequencing on the BGIseq-500 platform. Quality controls (QCs) on RNA samples (RIN/RQN ≥ 7.0, 28S/18S ≥ 1.0) were confirmed by BGI, followed by globin mRNA removal and cDNA library construction. Clean data of at least 4Gb (20M clean reads) per sample were collected.

After QC of fastq files, an mRNA-seq count table was obtained from bam files. Gene expression analysis was done on NetworkAnalyst platform using DESeq2 (*31*). Counts with variance percentile rank < 15% and counts < 4 were filtered out, transformed and normalized to Log2 reads per million values. Fold changes (Log2FC) were calculated for differentially expressed genes (DEGs) between FES patients and HCs, with significance of *p* < 0.05 adjusted by Benjamini-Hochberg (BH)’s false discovery rate (FDR). Monocyte-specific DEGs according to the relevant literatures of monocytic transcriptomic profiling (*14, 17, 32, 33*) were further selected for subsequent analysis.

Monocytic DEGs were subjected to analyses of gene ontology (GO)-based biological processes (BP), Kyoto Encyclopedia of Genes and Genomes (KEGG), and protein-protein interaction (PPI) by Search Tool for the Retrieval of Interacting Genes (STRING), with FDR < 0.05 as the cut-off for significantly enriched GO terms and KEGG pathways. PPI network was set at a high confidence threshold of 0.7 and clustered with k-means method. Plots of DEGs were made using R, GraphPad, and online Morpheus.

### Blood sampling and flow cytometric staining of peripheral blood monocytes

Five (5) ml of fresh heparin lithium-anticoagulated peripheral blood samples were collected from 29 FES patients and 27 HC subjects of the above participants after overnight fast, and processed within half an hour for fluorescent staining of cell surface receptors as described previously (*27*). The fluorochrome-conjugated antibodies used in this study were 10 μl FITC-labeled mouse anti-human CD14 (Clone M5E2; Catalog Number 555397; BD Biosciences) and 3 μl PerCP-Cy™5.5-labeled mouse anti-human CD16 (Clone B73.1; Catalog Number 565421; BD Biosciences). The percentages of classical (CD14++CD16−), intermediate (CD14++CD16+) and non-classical (CD14+CD16++) subsets among the total monocyte population were determined based on corresponding gatings. Samples were acquired by a BD FACSCalibur flow cytometer and the analyses were performed with FlowJo V10 software, as shown in Fig. 2A.

**Fig. 2.**
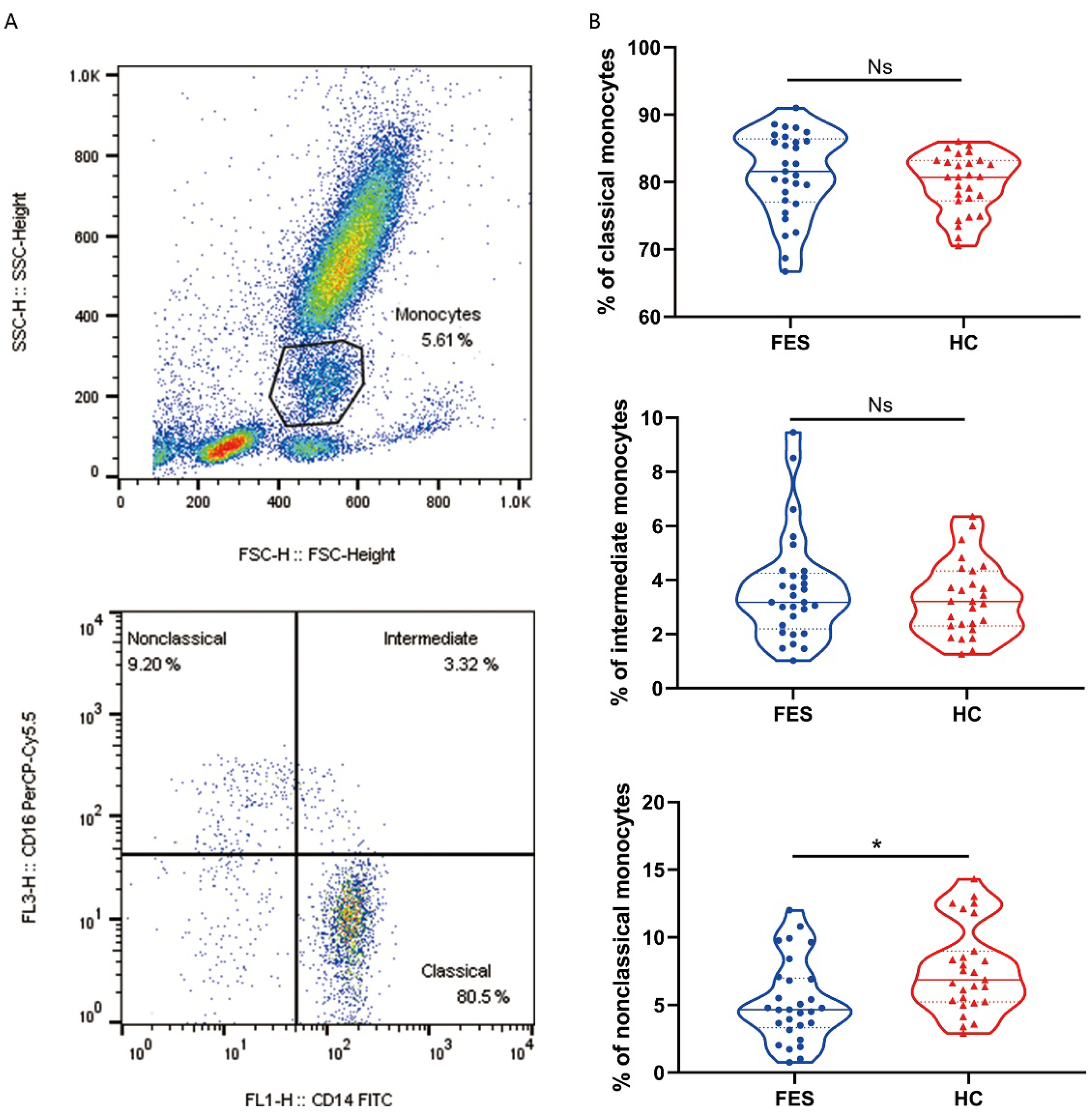
Nonclassical monocytes are decreased in the peripheral blood of FES patients. Whole blood sample was stained with monocytic markers FITC-labeled mouse anti-human CD14 and PerCP-Cy™5.5-labeled mouse anti-human CD16 and measured by flow cytometry. **(A)** Dotplots show gating strategy. Monocytes were defined according to surface expressions of CD16 and CD14. The CD14++CD16− classical monocytes, CD14++CD16+ intermediate monocytes, and CD14+CD16++ nonclassical monocytes were quantitated, respectively. **(B)** Violin plots show the blood monocytic subsets in FES patients and HCs. Solid and dotted lines represent the median and upper/lower quartile values, respectively. Ns, Not significant. *FDR < 0.05.

### Psychopathological symptoms and cognitive assessments

Psychopathological symptoms of the patients were assessed using the Positive and Negative Syndrome Scale (PANSS) by one of two senior psychiatrists who received a training session simultaneously prior to the study initiation for the consistency and reliability of ratings. Their intra-class correlation coefficient (ICC) reached above 0.8 for the PANSS total scores.

The MATRICS™ Consensus Cognitive Battery (MCCB) test was applied to assess the cognitive functioning for the subjects. It consists of ten tests encompassing seven cognitive domains, and domain scores as well as a composite score were computed using the MCCB scoring program. The clinical validity and reliability of Chinese version of MCCB had been previously established in both healthy volunteers and schizophrenia patients (*34*).

### Magnetic resonance image (MRI) acquisition and processing

Structural T1-weighted MRI brain images were acquired with a 3.0-T Prisma scanner (Siemens, Germany) and a 64-channel head coil at the Brain Imaging Center of Beijing Huilongguan Hospital. We used a sagittal 3D magnetization-prepared rapid acquisition gradient echo (MP-RAGE) sequence to get the T1 structural images. The acquisition parameters were as follows: repetition time (TR) =2,530 ms, echo time (TE) =2.98 ms, field-of-view (FOV) =256×224 mm, flip angle (FA) =7°, matrix size =256×224, thickness/gap =1/0 mm, and inversion time (TI) =1100 ms. Cortical thickness was estimated using FreeSurfer v5.3 (http://surfer.nmr.mgh.harvard.edu/) following the protocol of the Enhancing Neuro Imaging Genetics Through Meta Analysis (ENIGMA) Consortium (http://enigma.ini.usc.edu/). Images were visually checked for quality to make sure that cortical regions were properly segmented and labeled. Thirty-four distinct gyral-defined regions per hemisphere were extracted according to the Desikan-Killiany atlas (*35*). All of the cortex regions were averaged for right and left hemispheres (*36*). Brain images were obtained within 7 days after signing informed consent form.

### Statistical methods

Statistical procedures were performed with the IBM SPSS Statistics 21.0, and all missing values were treated as missing data. Normality of distribution was confirmed through the Shapiro-Wilkinson test. Demographic and clinical data were compared between the two groups by Student’s *t*-test, Mann-Whitney *U* test, or Chi-Square test. Pearson and Spearman correlation were used to determine associations between variables where appropriate. Age and sex were used as covariates for ANCOVA as well as partial correlation analyses. For analysis involving cognitive function, the covariates also included the education years. Partial correlations were employed to test the relationships among normalized RNAseq counts of monocytic DEGs, cortical thicknesses, and cognitions. Two-tailed *p* values were corrected for multiple testing with the BH’s FDR < 0.05 considered as significance. The PROCESS v3.3 for SPSS was used to perform the mediation analyses, with age, gender, and education years as covariates. The 95% confidence intervals (CIs) of direct and indirect effects were obtained from 5000 bootstrap samples. Statistically significant mediation effect was determined if the CI of indirect path did not contain zero.

## RESULTS

### Demographic and clinical data

A total of 128 FES patients and 111 healthy subjects were recruited according to the inclusion and exclusion criteria. No difference was observed between the groups for age, sex and smoking status (all *p* > 0.05) (Table 1). Years of education was significantly less in FES patients than that in HCs (*p* < 0.01) (Table 1).

**Table 1.**
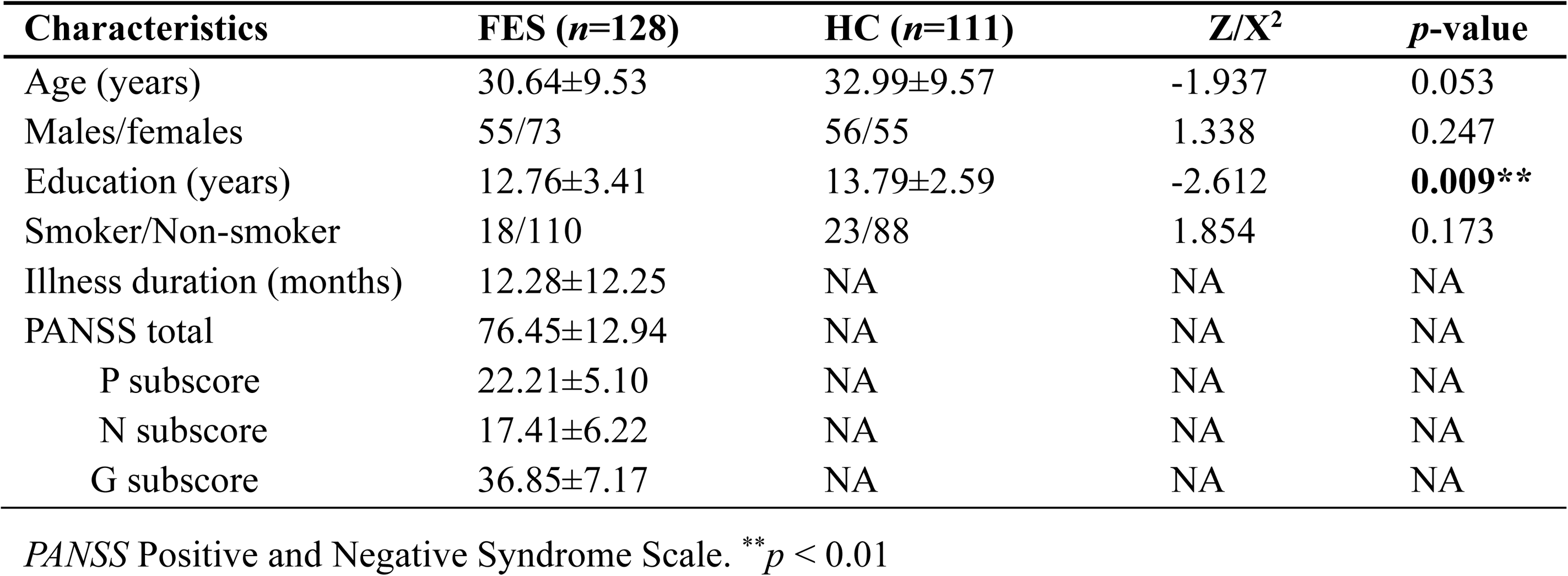
Participant demographic and clinical characteristics.

In the patient group, 18 patients were drug-naïve and 110 patients had been exposed to antipsychotics for 1-14 days (median 4 days) at the time of blood drawing. The majority of patients were taking risperidone (n = 48), aripiprazole (n = 16), risperidone combined with haloperidol injection (n = 15), and olanzapine (n = 14) (Supplementary Table 1).

### Peripheral blood RNAseq transcriptomic profiling for monocytic genes

We first studied the molecular signatures of circulating leukocytes and identified 9062 DEGs (FDR < 0.05) between FES patients and HCs, including 4479 upregulated and 4583 downregulated genes (Fig. 1A). To get a better insight into the gene expression changes among CD14/CD16-subsets of monocytes in schizophrenia, 79 subset-specific signature genes were chosen based on recent monocytic transcriptomic profiling work (*14, 17, 32, 33*), among which 54 were found to have FDR < 0.05 in our RNAseq datasets (Fig. 1A-1C, Supplementary Table 2). These 54 DEGs were enriched in different monocytic subtypes, respectively 4 (7.4%), 9 (16.7%), 20 (37.0%), and 21 (38.9%) belonging to all monocytes, classical, intermediate and nonclassical monocytes (Fig. 1B). In addition, among these subclasses, the majority (6 out of 9, i.e. 66.7%) of DEGs belonging to the classical monocytes were downregulated and that (13 out 21, 61.9%) of DEGs belonging to nonclassical monocytes were upregulated, while the proportions of up- and down-regulated genes (45% *vs.* 55%) were more equal in intermediate monocytes (Fig. 1B). Notably, among the DEGs with the most significantly changed expression (|Log2FC| > 0.5), S100 Calcium Binding Protein A family genes (*S100A8, S100A9* and *S100A12*, all downregulated) belonging to intermediate monocytes and Interferon Induced Transmembrane Protein family genes (*IFITM2* and *IFITM3*, both upregulated) belonging to nonclassical monocytes outstood (Fig. 1C).

**Fig. 1.**
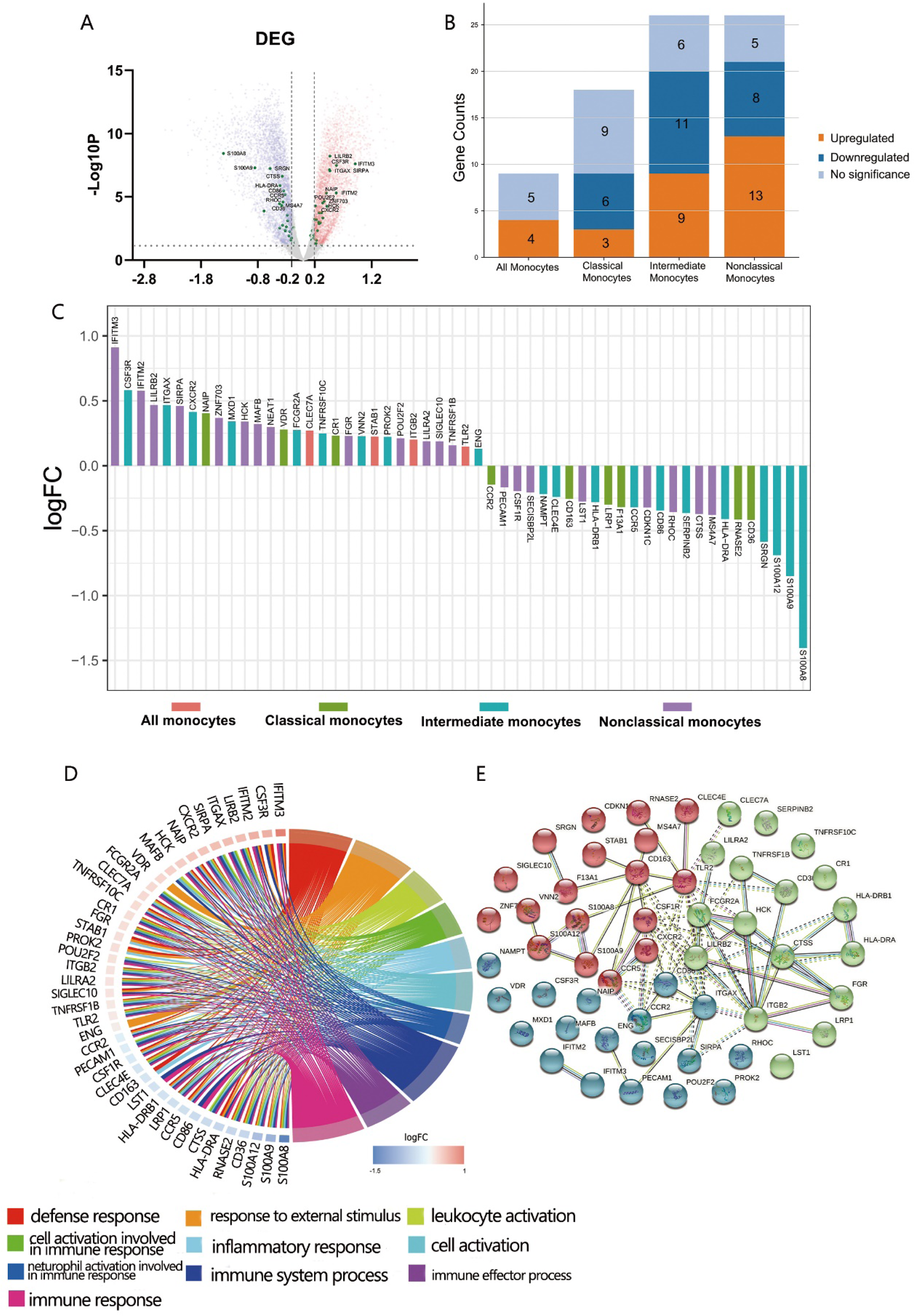
Peripheral blood RNAseq transcriptomic profiling shows differentially expressed monocytic genes between FES patients and HCs. **(A)** Volcano plot indicates the DEGs (FDR < 0.05, Log2FC > |0.2|) depicted as upregulated (red) or downregulated (blue). Fifty-four monocyte subset-specific DEGs are highlighted (green) and those with −Log10*P* > 3 are nominated. **(B-C)** These DEGs (FDR < 0.05) were especially enriched in intermediate and nonclassical monocytes. **(D)** Chord plot shows top 10 overrepresented GO-BP subontology for the 54 DEGs associated with monocytes. Genes are ordered according to the observed logFC and linked to their assigned terms via colored ribbons. **(E)** PPI analysis shows the interactions of DEGs set at a high confidence threshold of 0.7 and clustered with k-means = 3. Line between nodes features the type/strength of an interaction according to annotations in String.

We next checked the functions of these DEGs based on GO terms. From the 346 overrepresented GO terms of Biological Processes (FDR < 0.05) associated with them, expectantly the top ten ranked were mainly involved in immune and inflammatory response, with the largest set of DEGs enriched in immune system process, followed by immune response, defense response and response to external stimulus (Fig. 1D). Similarly, KEGG analysis revealed that the most significantly enriched pathways were infectious and autoimmune diseases (Supplementary Table 3). It is noteworthy that major histocompatibility complex, class II, DR alpha and beta 1 (*HLA-DRA/B1*) were observed in the majority of the KEGG enriched pathways.

PPI analysis of these monocytic DEGs further revealed a network containing 53 nodes and 75 edges, with PPI enrichment *p*-value < 1.0×10^-16^. Under the clustering criterium of k-means = 3, the network can be further grouped into the three interconnected clusters, in which genes such as the *S100A* family members, the *IFITM* family members, and the *CR1*-*ITG* family members are covered, respectively (Fig. 1E).

### Comparisons on blood monocytic subsets between FES and HC

A subgroup consisted of 29 FES patients and 27 HCs, of whom 23 FES patients and 26 HCs were also included in the total cohort, was used to explore the distribution of monocytic subsets by flow cytometry. There were no significant differences in age, sex, education years, or smoking status between FES and HCs in this subgroup, and their clinical characteristics are displayed in Supplementary Table 4A–4B.

Interestingly, FES patients had a decreased percentage of nonclassical monocytes compared to HCs after controlling for age and sex (5.27 ± 3.01% *vs.* 7.61 ± 3.21%, F = 8.736, FDR = 0.015) (Fig. 2B). We found no group differences in the percentage of classical and intermediate monocytes. Besides, the two groups did not differ in neither the absolute count nor the percentage of total blood monocytes according to the routine clinical blood test (all *p* > 0.05).

### Comparisons on brain cortical thickness and cognition between FES and HC

A subset of 60 FES patients and 54 HCs (two patients and two controls did not complete the MCCB assessment) from the total cohort, including the participants in the above flow cytometric experiment, underwent further MRI and MCCB tests. There were no significant differences in age, sex, education years, or smoking status between FES and HCs in this subset, and their clinical characteristics are displayed in Supplementary Table 4C–4D.

On the structural imaging aspect, the whole-brain average cortical thickness was reduced in FES patients versus HCs after controlling for age and sex (2.54 ± 0.08 vs. 2.59 ± 0.09, F = 10.078, *p* = 1.946×10^-3^). As illustrated in exemplary MRI images in Fig. 3A, among the 34 cortical regions defined by the DK atlas, 8 regions, i.e. supramarginal gyrus, inferior parietal cortex, superior parietal gyrus, lateral occipital cortex, inferior temporal gyrus, precuneus, fusiform gyrus, and superior temporal gyrus were all remarkably thinner in FES individuals, while their pericalcarine and lingual gyrus were statistically thicker than those of HCs (all FDR < 0.05) (Supplementary Table 5 and Fig. 3B).

**Fig. 3.**
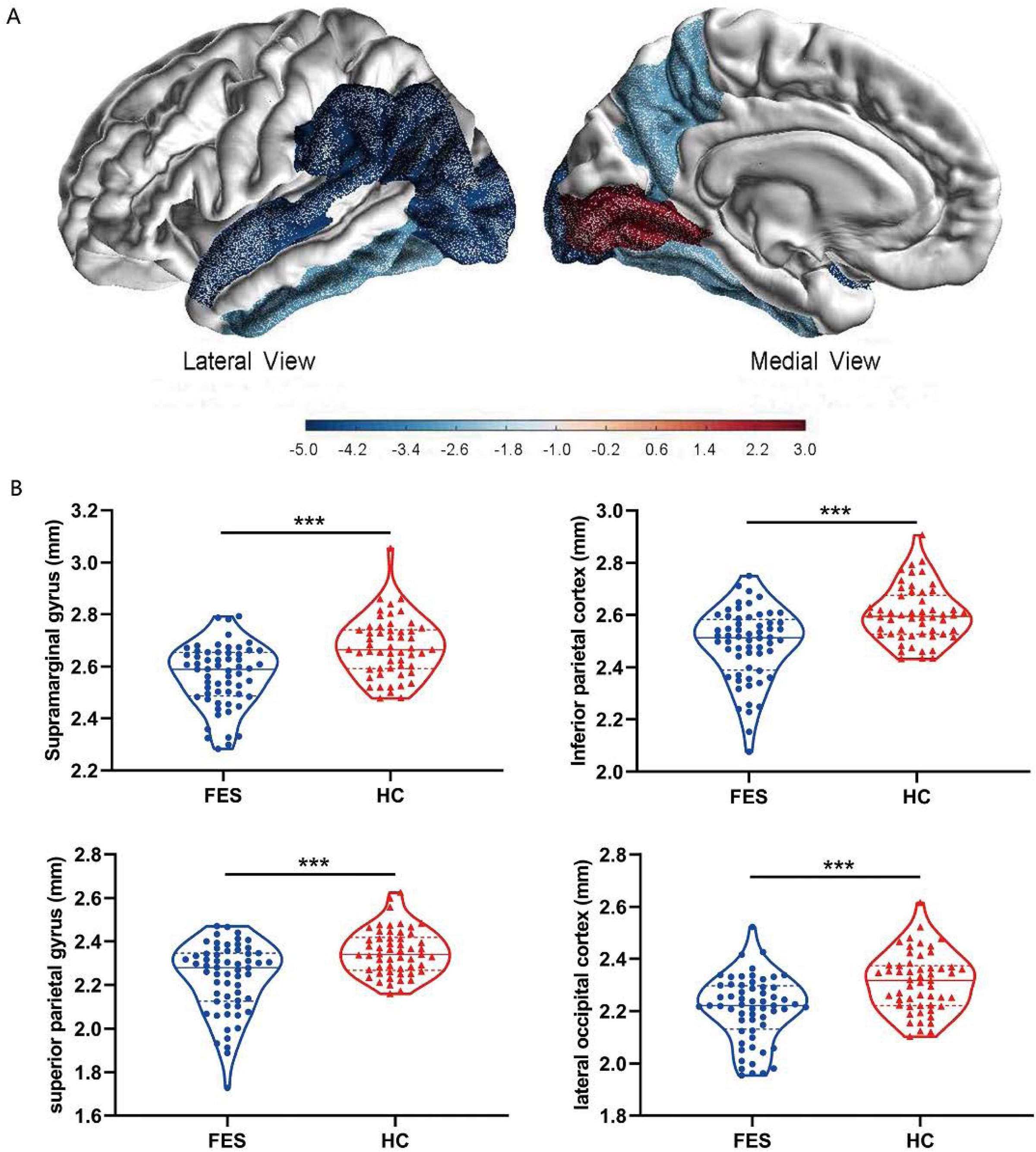
Brain cortical thicknesses are different between FES patients and HCs. **(A)** Thirty-four distinct gyrus-defined regions per hemisphere were extracted according to the Desikan-Killiany atlas and averaged for both hemispheres. Color coding in the exemplary MRI images represents the significance (t value) in difference of cortical thickness between FES patients and HCs, after FDR correction controlling for age and sex. **(B)** Brain regions with the most significant reduction in cortical thickness of FES patients are shown, with median and upper or lower quartile drawn in violin plots, respectively. ***FDR < 0.001.

As expected for MCCB test, FES patients also showed significantly lower total MCCB score and the seven domain subscores compared to HCs after controlling for age, sex, and education years (all FDR < 0.001) (Table 2).

**Table 2.**
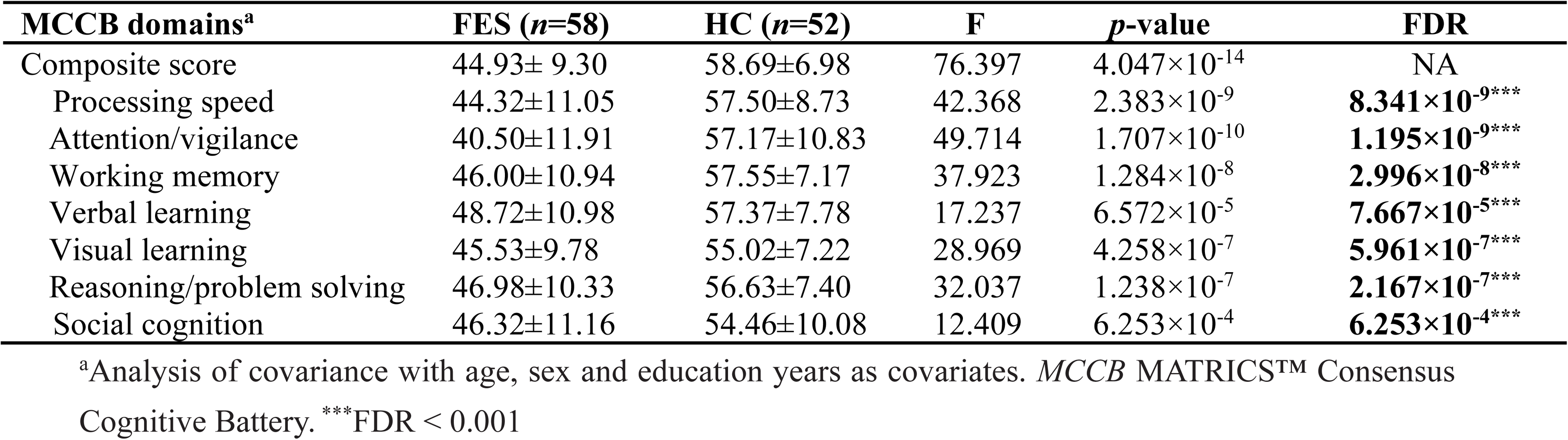
Comparison of the MCCB scores between first-episode schizophrenia patients and healthy controls.

### Associations among monocytic subset signature genes, cortical thickness, and cognition

To explore the relationships of monocytic DEGs with brain structure and cognition, we performed partial correlational analyses of the 54 DEGs RNAseq counts, the thicknesses of the 34 brain cortical regions and the MCCB scores, controlled for age, sex, and education years in the FES and HC groups, respectively (Fig. 4). Clustered heatmaps of the correlational matrices of these three modules showed that the main differences between the two groups included two aspects. Firstly, for the HCs, the internal associations among the 54 DEGs inside the gene module and the relationships among the 34 brain regions were both dominated by positive correlations (Fig. 4A). Nevertheless, these internal correlations were both attenuated in the FES group, especially the interior correlations between genes related to the nonclassical monocytes and between the encephalic regions associated with the occipital lobe (Fig. 4B). Secondly, as a whole profile, monocytic DEGs expressions were in a strikingly inversive relationships with both cortical thicknesses and MCCB scores in the HC group (Fig. 4A). However, such negative relationships were both weakened in the FES patient group (Fig. 4B).

**Fig. 4.**
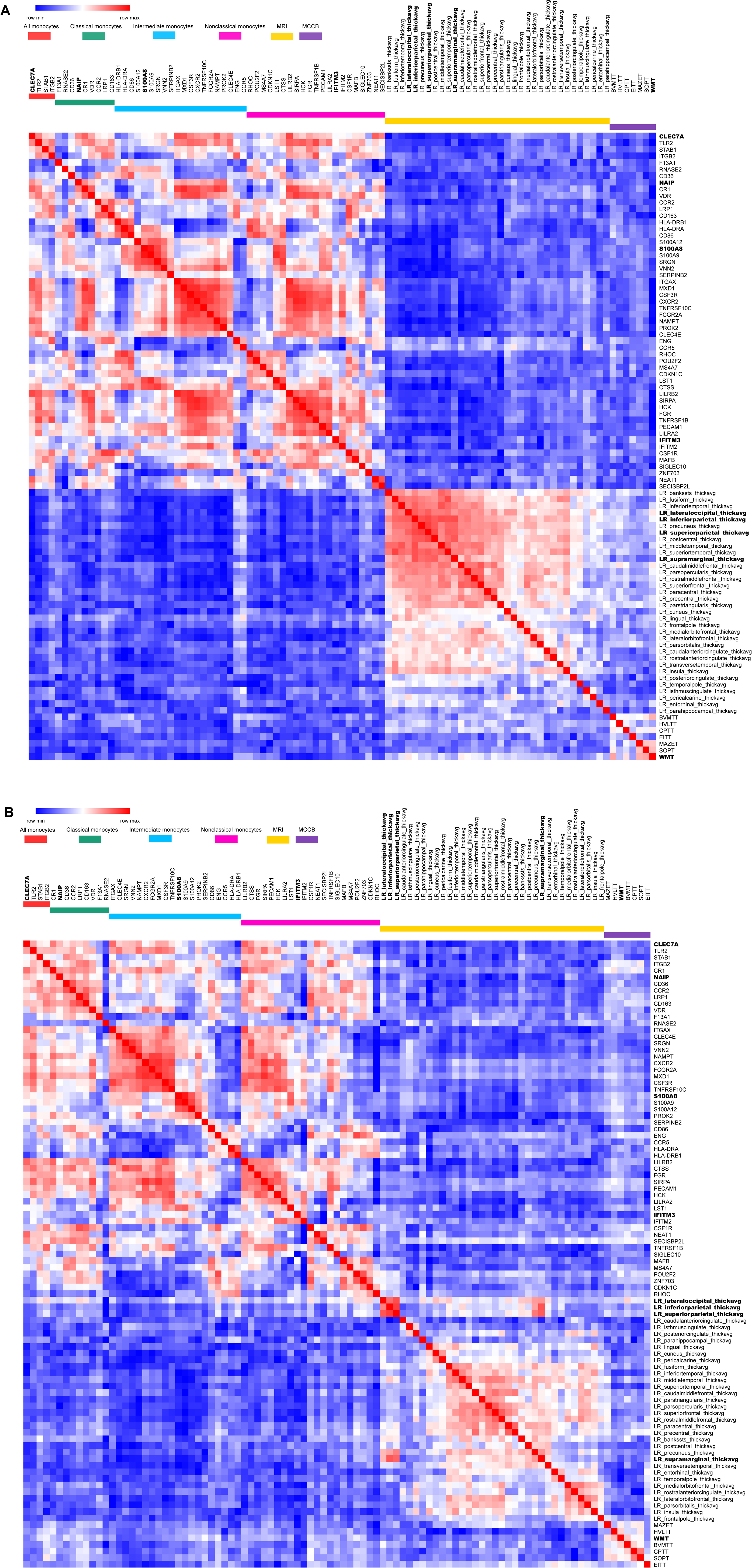
Clustered heatmaps of correlational matrices among monocytic DEGs expression, cortical thicknesses, and cognition show negative impacts of genes on cortex and cognition and differences between FES patients and HCs. Partial correlational analyses of the 54 monocytic DEGs normalized RNAseq counts, the thicknesses of the 34 brain cortical regions and the MCCB scores, controlled for age, sex, and education years in HCs **(A)** and FES patients **(B)**, respectively. Heatmap colors represent the negative (blue) and positive (red) partial correlation coefficient, respectively. Monocyte subsets, cortical regions and MCCB tests are categorized with respective color bars, and variables showing the most significant difference between FES patients and HCs in each category are in bold.

Next, we checked the detailed correlations between DEGs and each cortical region, controlling for age, sex, and education years and setting the significant level at *p* < 0.01, as visualized in volcano plots (Fig. 5). In the HC group, numerous negative correlations appeared, with the top outstanding ones coming from genes that were all upregulated in FES patients compared to HCs (Fig. 1C), namely a pan-monocytic gene C-Type Lectin Domain Containing 7A (*CLEC7A*), a classical monocytic gene NLR family Apoptosis Inhibitory Protein (*NAIP*) and an intermediate monocytic gene Prokineticin 2 (*PROK2*) (Fig. 5A). In the FES group, the most highly significant correlation came from a classical monocytic gene Ribonuclease A Family Member 2 (*RNASE2*), which was downregulated in FES patients compared to HCs (Log2FC = −0.41, FDR = 0.003, Supplementary Table 2) and was inversely correlated with the lateral occipital cortical thickness (Fig. 5B). Besides, the intermediate monocytic genes of the S100A family members were inversely associated with the thicknesses of the precentral, middle temporal and paracentral cortices in the FES group (Fig. 5B). Among all the correlations in both groups, only the negative relationship between the *RNASE2* and the thickness of the lateral occipital cortex in the patient group survived BH correction for 54×34 tests (partial *r* = −0.531, FDR = 0.040).

**Fig. 5.**
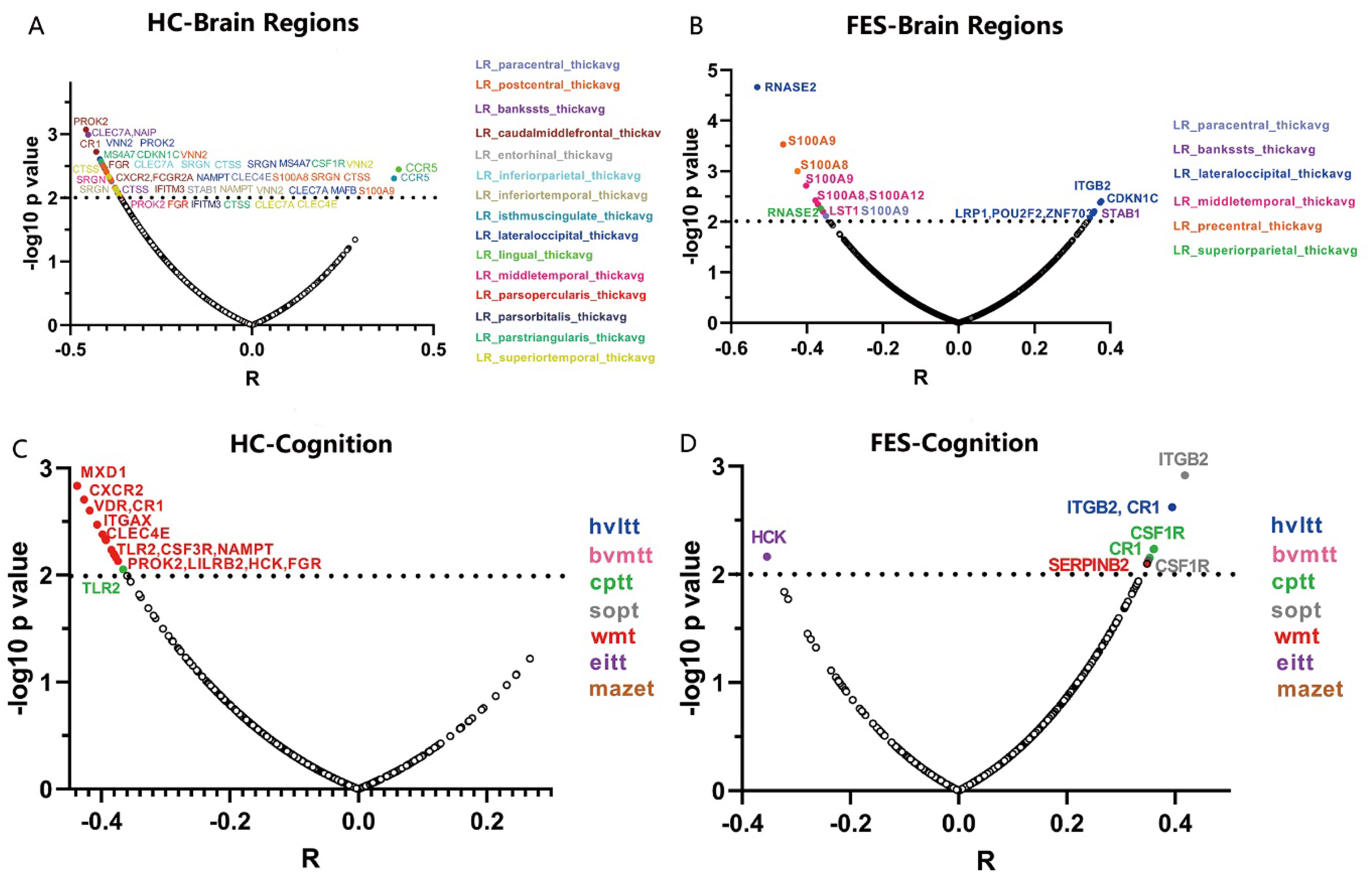
Correlations of monocytic DEGs mRNA levels with cortical thicknesses and MCCB scores show more extensive negative impacts of the genes in HCs than FES patients. **(A-D)** Partial correlational analyses between 54 monocytic DEGs and 34 cortical regions controlling for age, sex, and education years in HC **(A)** and FES **(B)** groups, respectively; and with 7 MCCB subscale scores in HC **(C)** and FES **(D)** groups, respectively. The abscissa represents partial correlation coefficient. The significant level was set at *p* < 0.01. Among all the correlations, only the inverse relationship between the classical monocytic gene *RNASE2* and lateral occipital cortical thickness in FES patients passed the FDR (54×34) correction.

In addition, we also explored whether the percentage of nonclassical monocytes was associated with the cortical thickness. The results showed that among FES patients, after adjustment for age, sex, and education years, the percentage of nonclassical monocytes was inversely correlated with the cortical thicknesses of seven anatomic regions, including the paracentral lobule, entorhinal cortex, fusiform gyrus, middle temporal gyrus, pars opercularis of inferior frontal gyrus, superior temporal gyrus, and temporal pole (partial *r* within the range of −0.5 ~ −0.7, all FDR < 0.05) (Supplementary Fig. 1A). No statistically significant relationships between the percentage of nonclassical monocytes and the thicknesses of cortical regions were detected in the HC group (all FDR > 0.05, Supplementary Fig. 1B).

Also, correlations between DEGs and cognitive functions revealed negative associations between expressions of a variety of monocytic genes and MCCB scores in the HC group (Fig. 5C), among which some were reverted to positive relationships in the FES group, especially the complement C3b/C4b receptor 1 (*CR1*), integrin family (*ITG*), and colony stimulating factor receptor family (*CSFR*) genes that belong to classical-intermediate monocytes (Fig. 5D), but none of these correlations passed the FDR correction. Besides, there were no significant correlations between the percentage of nonclassical monocytes and all the MCCB domains in both groups even at nominal levels (all *p* > 0.05).

### Mediation effect of the cortical thickness on the association of monocytic gene and cognition

Finally, considering the overt relationship between the gene *RNASE2* and the lateral occipital cortical thickness in the FES group, we further explored whether the lateral occipital cortex may mediate the *RNASE2*-cognition relationship in patients. Because the thickness of the lateral occipital cortex was only associated with the BVMT of MCCB at nominal level (partial *r* = 0.370, nominal *p* = 0.005), BVMT score was used as the dependent variable in the mediation analysis. Introducing the lateral occipital cortical thickness as a mediator, the indirect path (path ab) from expression level of *RNASE2* to BVMT was significant (β = −0.156, 95% CI, −0.359 to −0.003), while the direct effect (path c’) between them was insignificant (β = −0.047, t = −0.367, *p* = 0.715), which implied that the effect of *RNASE2* on BVMT score was fully mediated by the lateral occipital cortex (Fig. 6).

**Fig. 6.**
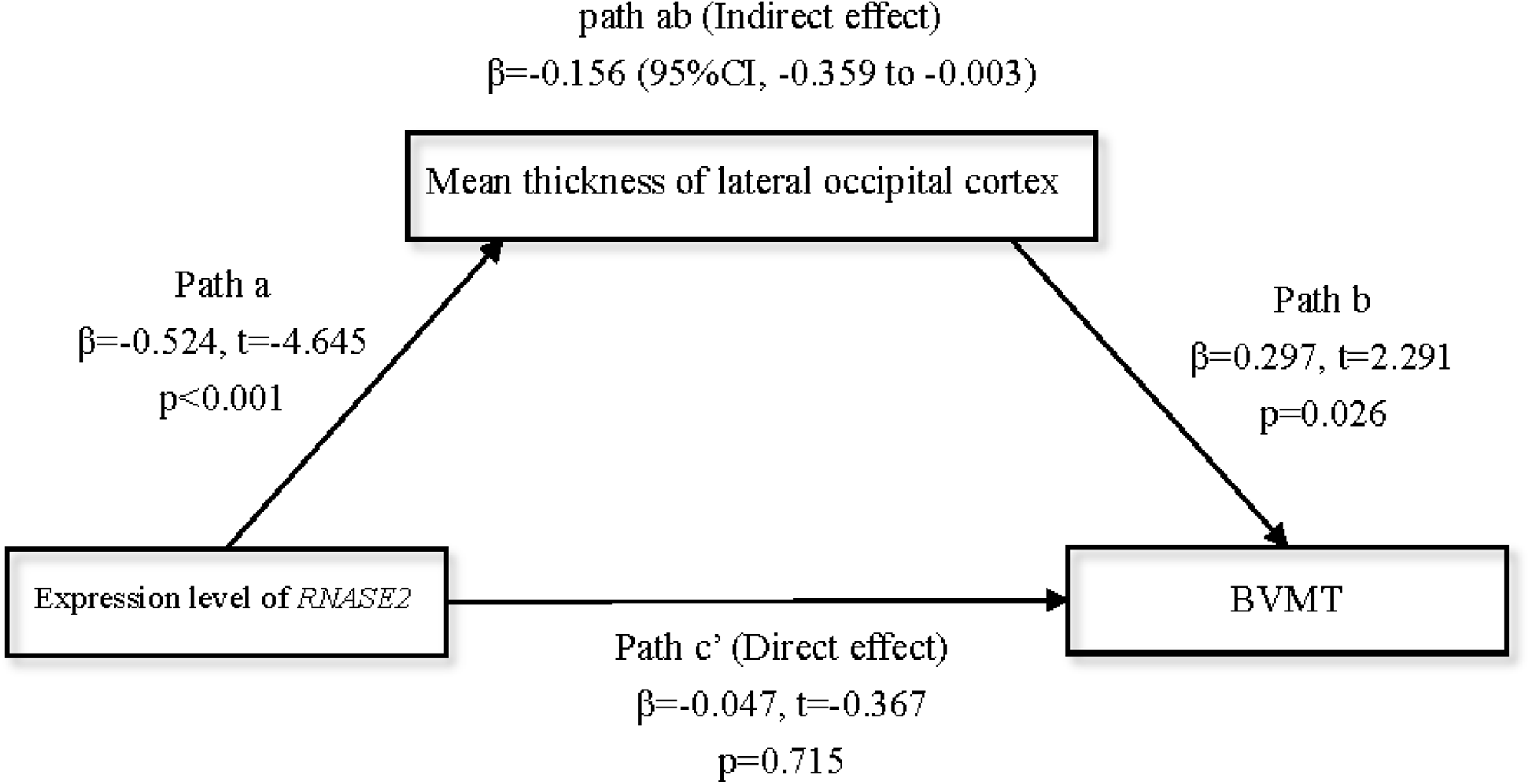
Full mediation effect of the lateral occipital cortex between the gene RNASE2 and visual memory. X = *RNASE2* expression level; Y = Brief Visuos-patial Memory Test (BVMT) score; Mediator = lateral occipital cortex. Age, sex, and education years were included as covariates.

### Association with clinical psychiatric symptoms

None of the 54 DEGs expressions and the cortical thicknesses of 34 brain regions were significantly correlated with PANSS positive, negative, and general psychopathology symptom scale scores, respectively, after FDR correction (all FDR > 0.05), although the correlation heatmap showed that the relationship between the modules of cortical regions and PANSS were dominated by negative correlations (Supplementary Fig. 2A).

Additionally, the percentage of intermediate monocytes was inversely associated with the PANSS negative symptoms (*r* = −0.480, *p* = 0.008) (Supplementary Fig. 2B), whereas no other correlations between monocytic subsets and PANSS subscales scores were observed (all *p* > 0.05).

## DISCUSSION

Previous studies have not considered monocytes as a highly heterogeneous population that may have complicated roles in schizophrenia, thus, we sought to scrutinize them in more details. The major findings in our present study are (1) the monocyte-related transcriptomic profiles showed significantly changed expressions in an early course of schizophrenia onset, especially for intermediate and nonclassical monocytic subsets, with the most outstanding alterations being downregulated *S100A* and upregulated *IFITM* family members belonging to the intermediate and nonclassical monocytes, respectively; (2) the inverse inter-relationships of monocytic DEGs with cortical thicknesses and cognition, which were weakened or even reversed in FES patients compared to HCs; while the percentage of nonclassical monocytes, which was decreased in schizophrenia, was negatively associated with the thicknesses of multiple brain cortical regions in FES patients; (3) the lateral occipital cortex fully mediated the negative effect of the classical monocytic gene *RNASE2*, a gene encoding an eosinophil-derived neurotoxin, on visual learning and memory in FES patients. An overall impression on these results was that monocytic subsets and their genes may play a differentially detrimental role for brain and cognition in healthy people, but FES patients may have developed a to-be-defined immune/hematopoietic mechanism to compensate this effect.

### Decreased proportion of nonclassical monocytes and upregulated nonclassical monocytic DEGs in schizophrenia

We studied peripheral blood by flow cytometry and RNAseq to profile monocytic functions in FES, providing the first evidence to our knowledge that monocytic defects may be implicated in the pathogenesis at early stage of schizophrenia. Recent transcriptomic studies using blood of FES patients, postmortem brains, and patient-derived cerebral organoids generated from induced pluripotent stem cells have also demonstrated dysregulation of inflammation and immune function in this disease (*37–39*). To scrutinize specific monocytic functions with higher detail, we categorized the DEGs into different monocytic subtypes.

Decreased proportion of nonclassical monocytes along with upregulated signature genes in FES patients are the most interesting findings in the present study, although the exact underlying mechanism awaits to be revealed. We found that more DEGs fell into the intermediate and nonclassical monocytes compared to the classical subset in patients, suggesting that biological functions of these two subpopulations may be more affected. We speculate that the aberrant transcriptomics and the looser inter-correlations among the DEGs associated with monocytic subsets in patients indicate an imbalanced monocytic differentiation process, possibly due to abnormal hematopoiesis/myelogenesis or activation of mature monocytes by infections in FES patients. Moreover, nonclassical monocytes play an important role in maintaining vascular homeostasis (*18*) and studies have demonstrated that patients with schizophrenia had vascular endothelial pathologies due to chronic low-grade systemic inflammation (*40, 41*). This possibly leads to nonclassical monocytes recruitment to repair vascular endothelium, resulting in nonclassical monocytopenia in the peripheral blood. This is similar to an observed phenomenon in patients with severe forms of lupus nephritis, showing that lower levels of nonclassical monocytes in the peripheral blood was accompanied by a higher degree of infiltrates of CD16+ cells in the glomerulus (*42*). We also found inverse correlations of the percentage of blood nonclassical monocytes with the thicknesses of 7 brain cortical regions, with 5 belonging to the temporal lobe, in patients but not healthy participants. Since the temporal cortex regulates the subcortical limbic structure and the hypothalamic– pituitary–adrenal axis that are known to in turn regulate immune cell mobilization in the circulation (*43*), the cortical dystrophy in FES patients may also explain the reduced amount of blood nonclassical monocytes in our study. Nevertheless, our study may be affected by its cross-sectional nature and the small sample size, and these speculative accounts deserve further investigation.

Interestingly, among the DEGs belonging to classical monocytes, most were downregulated in FES patients. Likewise, in intermediate monocytes, downregulations of the myeloid alarmin-related *S100A* genes (*S100A8*, *S100A9* and *S100A12*) in patients were the most notable changes. *S100A* family acts as damage-associated molecular patterns (DAMPs) on pattern recognition receptors, such as RAGE and TLR4 receptors, which are important for phagocytosis, cytokine release, cell adhesion and migration (*44, 45*). Previous studies have reported increased *S100A8*/*9*/*12* (S100A8 dimerizes with S100A9 to form calprotectin) gene expression in peripheral blood cells, prefrontal cortex, and hippocampus of individuals with schizophrenia (*38, 46–48*), which are inconsistent with our results, possibly due to the differences in disease staging and antipsychotics. In those studies, there were no restrictions on duration of disease and medication, whereas study by Santiago et al (*48*) suggested that enhanced *S100A8* gene expression may be a result of antipsychotic exposure. Our finding of significantly downregulated *S100A* family genes indicates the capacity of releasing DAMP signals from monocytes and other innate cells may be impaired.

Remarkably, our results showed upregulated *IFITM* family members belonging to nonclassical monocytes in FES patients, which is corroborated by earlier studies of the blood and postmortem brains of patients with schizophrenia, especially in the prefrontal cortex, hippocampus and cortical blood vessels, independent of antipsychotic use (*38, 49–53*). IFITM2/3 proteins, which are mainly localized to late endosomal/lysosomal membranes, have been shown to restrict a broad range of viral entry and replication, such as influenza A virus and cytomegalovirus (*54–56*), while Natalya et al. (*24*) found that the area and number of lysosomes of monocytes were significantly increased in schizophrenia patients as compared to HCs. Taken together with our findings on *IFITM2/3* genes, it is plausible that dormant or reminiscent viral infection may be associated with pathogenesis in FES, a hypothesis that has been persisted for decades and supported by epidemiological evidences on viral infection pandemics or perinatal immune activation models (*57–60*).

Taken together, our results highlight that not only global monocytic functions but also their subtypes may be more specifically defective in FES patients, which should not be neglected in this field of research.

### Associations among monocytic subset DEGs with cortical thickness and cognition

Our further finding was that FES patients exhibited widespread cortical thinning, primarily in the parietal, occipital and temporal cortices, in agreement with previous studies in FES patients (*61, 62*), suggesting that extensive cortical gray matter loss has taken place in the early phase of illness. However, unlike prior studies, we did not detect differences in the frontal and cingulate regions, possibly because we averaged the left and right hemispheres in our study.

Little is known about monocytic effect on the schizophrenic brain currently, but research findings in other fields may be enlightening. A recent study reported that higher plasma monocyte activation markers sCD14 and sCD163 were associated with smaller frontal and temporal cortical volumes among women with HIV (*63*). Moreover, chemokine receptor CCR2 on intermediate monocytes was correlated with disturbances of neuro-metabolites in the right and left caudate nucleus, contributing to HIV-associated neurocognitive disorders (*64*). Another related important research field is Alzheimer’s disease. Recently, disease-associated microglia implied in amyloidosis and cognitive impairment have been depicted in both animal model and humans of Alzheimer’s disease through single-cell RNAseq (*65, 66*). Several highly significant monocytic DEGs found in our current study, such as *CCR5*, *CLEC7A*, *CTSS*, *IFITM3*, and *ITGAX*, are associated with such microglial subtype. Also, a relationship between monocyte-derived inflammation and hippocampal atrophy as well as cognitive decline in Alzheimer’s disease had been reported (*67*). Intriguingly, with a data-driven approach here, we detected that the negative inter-relationships between monocytic DEGs and cortical thicknesses as well as cognition were weakened in schizophrenia patients as compared to healthy controls. These indicate to us that firstly, monocytic subsets in general negatively impact brain and cognition in normal people; secondly, disruption of this detrimental role in FES patients implies a functional loss or even compensatory mechanism of monocytes in the early stage of the disease.

Notably, among the DEGs associated with cortical thinning in both study groups, the classical monocytic gene *RNASE2* was the most striking one. The protein encoded by *RNASE2* is a cytotoxic protein that can induce proinflammatory cytokine production in monocytes/macrophages by acting as a DAMP (*68, 69*). It causes severe damage to myelins and loss of neurons in the rabbit brain, an event known as the Gordon phenomenon (*70, 71*). Possibly, a highly inverse association between *RNASE2* and lateral occipital cortical thickness in our FES patients may suggest that their thinner lateral occipital cortex may be particularly vulnerable to the neurotoxic effects of *RNASE2*, while the downregulation of *RNASE2* in patient group hints a self-protective response of host.

Of another note, among the negative associations with cortical thicknesses set at a less strict threshold (*p* < 0.01, uncorrected), intermediate monocytic genes of the *S100A* family members were more dominant in FES patients as compared to those of healthy individuals, implying *S100A* family members may also play a critical role in shaping cortical structure in schizophrenia, similarly as *RNASE2*.

The current study has several limitations that must be considered. Firstly, the cross-sectional design of this study limits our causal interpretations. Whether the perturbation of monocytic-transcriptome profiling and imbalanced distribution of monocyte subpopulations, along with their associations with cortical thickness and cognitive functions observed here are a nature of compensatory or causal effect remains unclear, although mediation analyses may indicate possible directionality. Secondly, we performed RNAseq by using the peripheral whole blood samples, hence, the transcriptomic changes in our study are not solely restricted to monocyte subsets, which should be confirmed by more advanced approach such as single-cell RNAseq. Thirdly, we did not validate the critical DEGs by employing quantitative-PCR, even though the current results could still offer a valuable preliminary transcriptional landscape of genes associated with monocyte subsets in the early course of schizophrenia. Fourthly, results from monocyte subtyping by flow cytometry should be interpreted with caution due to the small sample size in this research. Finally, for a better understanding of the complex bidirectional brain-immune communication, a larger sample of longitudinal design with information about early life exposure to immune damage, which could for instance induce trained innate immunity (*72*), will be required.

In summary, our findings demonstrate dysregulated transcriptomic profiles and disproportionality of monocyte subpopulations at early stage of schizophrenia, with intermediate and nonclassical subsets coupled with *S100A* and *IFITM* family members worthy of special attention. Some of these changes are linked to cortical structure, cognitive performances, and clinical symptoms, although some results did not survive multiple testing corrections. It is noteworthy that overall monocytic subsets negatively impact cortical thickness and cognition in both healthy subjects and schizophrenia patients. Furthermore, the lateral occipital cortex fully mediated the negative effect of a classical monocytic gene *RNASE2* on visual learning in FES patients. We have minimized antipsychotic treatment confounds, and these results suggest that deficits in monocytes may be vital for cognitive dysfunction of schizophrenia. Given the high heterogeneity of monocytes, a more spectral view instead of an ‘one model fits all’ scenario should be kept in mind in future schizophrenia research.

## Supplementary Materials

Fig. S1. The associations between the percentage of nonclassical monocytes and the thicknesses of cortical regions.

Fig. S2. Association with clinical psychiatric symptoms.

Table S1. Distribution of antipsychotics at the time of enrollment in the total cohort.

Table S2. 79 monocyte subset-enriched genes.

Table S3. KEGG pathway.

Table S4. Demographic and clinical characteristics in the participants receiving monocytic subtyping and brain imaging respectively.

Table S5. Brain regions with significant differences in cortical thickness between FES patients and HCs.

## Data Availability

Data and code are available from the authors upon reasonable request.

## Acknowledgements

We are grateful to all patients and healthy volunteers who participated in this research. We thank Ahto Salumets for helping on quality control and processing with RNAseq data.

## Funding

Academy of Finland research grant 273108 (LT) Estonian Research Council-European Union Regional Developmental Fund Mobilitas Pluss Program MOBTT77 (LT) National Natural Science Foundation of China 81761128021 (YLT) National Natural Science Foundation of China 82001415 (SC)

## Author contributions

The study was initiated and directed by LT and YLT. SC, MHX, PZ and TY were responsible of recruitment of study subjects and clinical assessments. FMF performed the imaging data analysis. LT, YLT, SC and KC performed statistical analysis and interpreted results. SC and LT wrote the first draft of the manuscript in consultation with YLT. KC, HZF, YMC, FDY, BPT and LEH were invited in evolving the ideas, analyzing data and editing the manuscript. All the authors read and approved the final manuscript.

## Competing interests

The authors declare that they have no conflict of interest.

## Data and materials availability

Data and code are available from the authors upon reasonable request.

---

**Supplementary Fig. 1.**
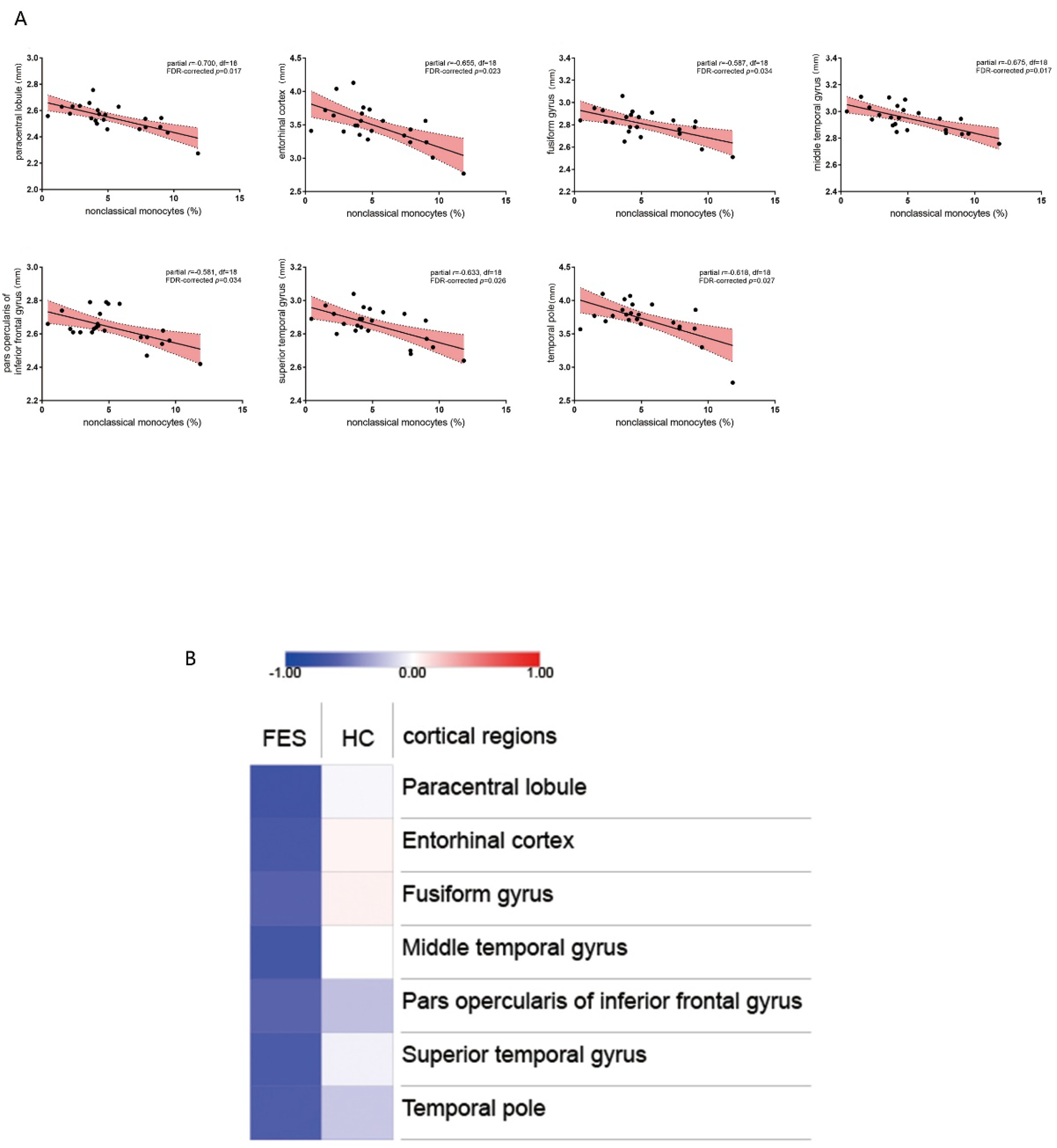
The associations between the percentage of nonclassical monocytes and the thicknesses of cortical regions. (**A**) Scatter plots indicated that the percentage of nonclassical monocytes was significantly inversely correlated with the cortical thicknesses of seven anatomic regions in FES group (all FDR < 0.05). partial r and p values were obtained after adjustment for age, sex and education years. Colored regions were 95% confidence bands. (**B**) Heatmap of correlations between the percentage of nonclassical monocytes associated with the seven cortical regions as above mentioned in both FES and HC groups. Color bar represents partial correlation coefficient.

**Supplementary Fig. 2.**
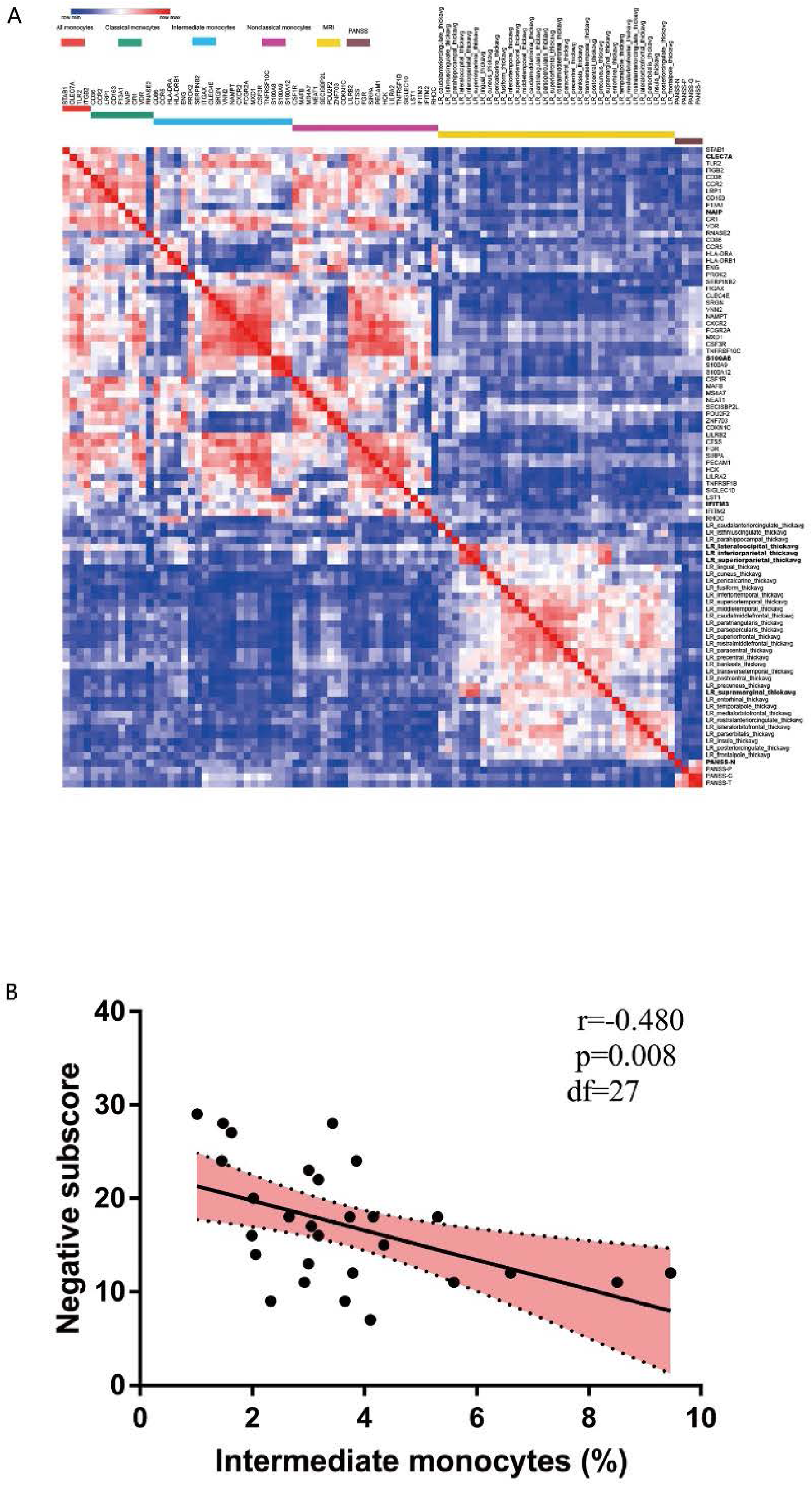
Association with clinical psychiatric symptoms. (**A**) Clustered heatmap of the correlational matrices among the 54 monocytic DEGs RNAseq counts, cortical thicknesses, and the PANSS scores in the FES group. Age, sex, and education years were controlled by partial correlational analyses. Heatmap colors represent negative (blue) and positive (red) coefficients, respectively. (**B**) Scatter plots indicated that the percentage of intermediate monocytes was inversely correlated with the PANSS negative symptoms (Spearman correlation). Colored regions were 95% confidence bands.

**Supplementary Table 1.**
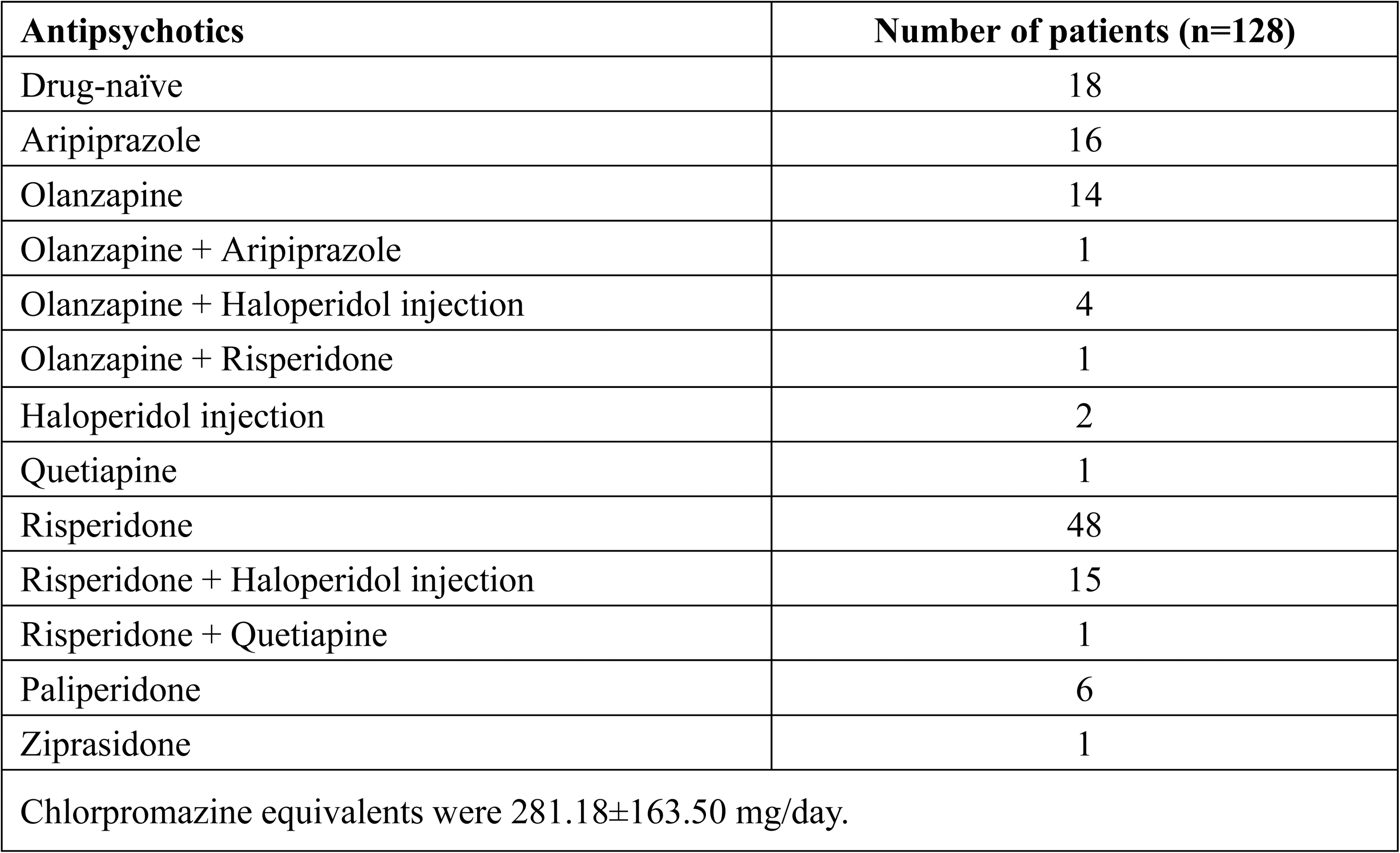
Distribution of antipsychotics at the time of enrollment in the total cohort.

**Supplementary Table 2.**
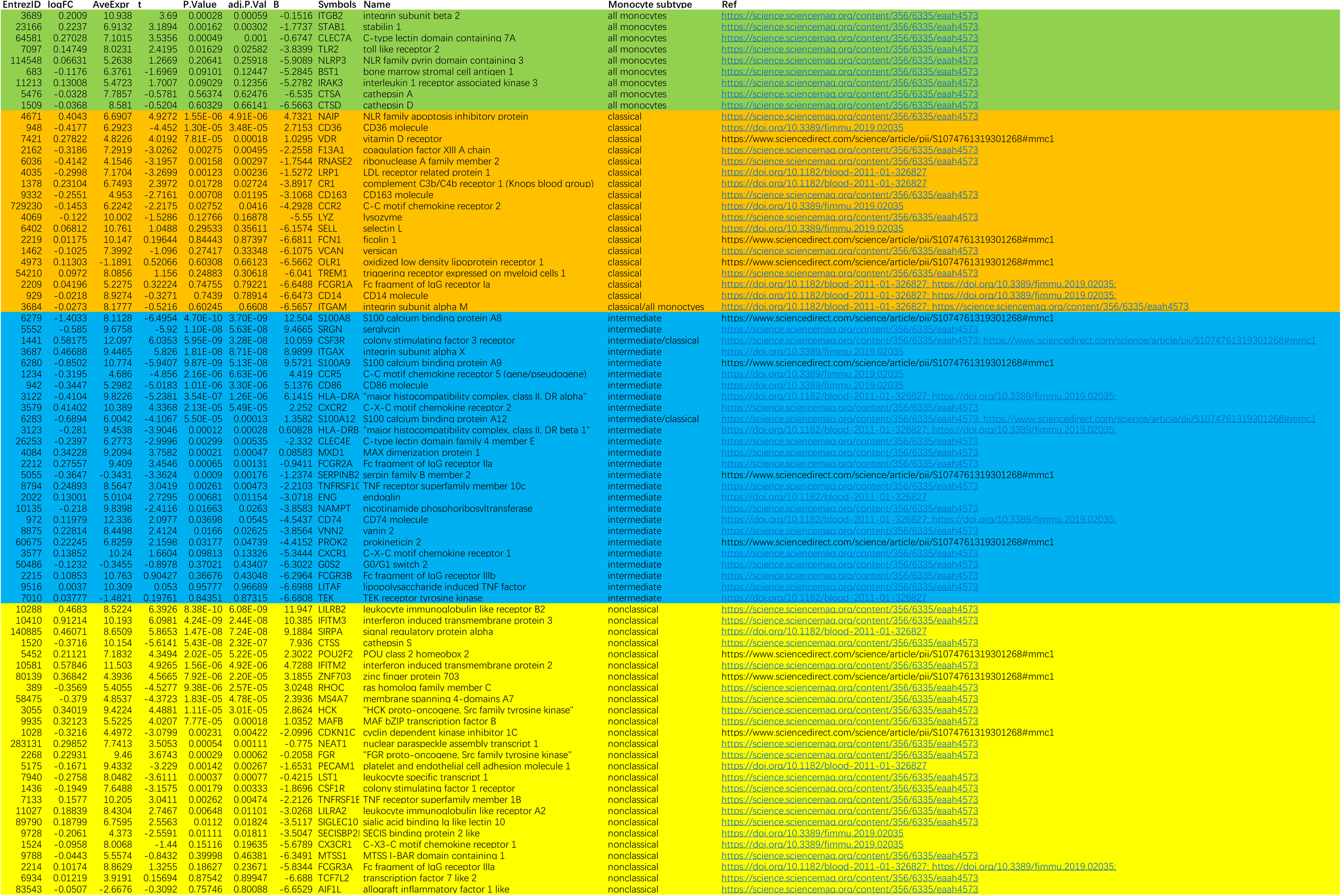
79 monocyte subset-enriched genes.

**Supplementary Table 3.**
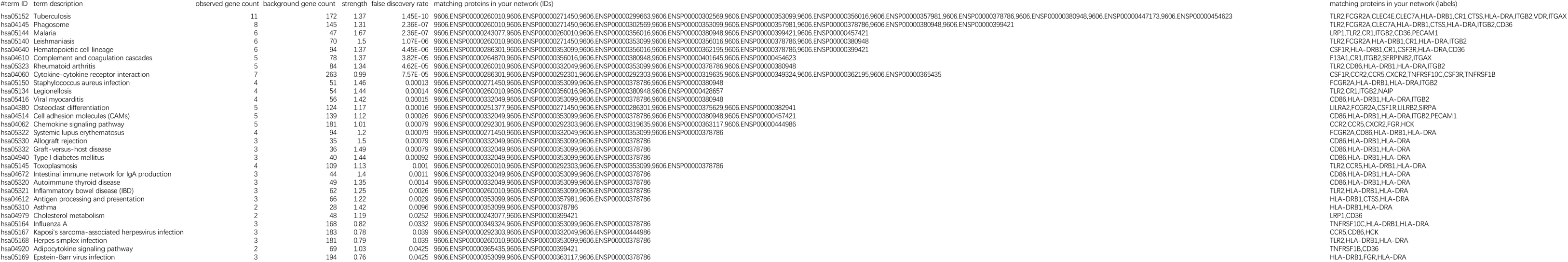
KEGG pathway.

**Supplementary Table 4A.**
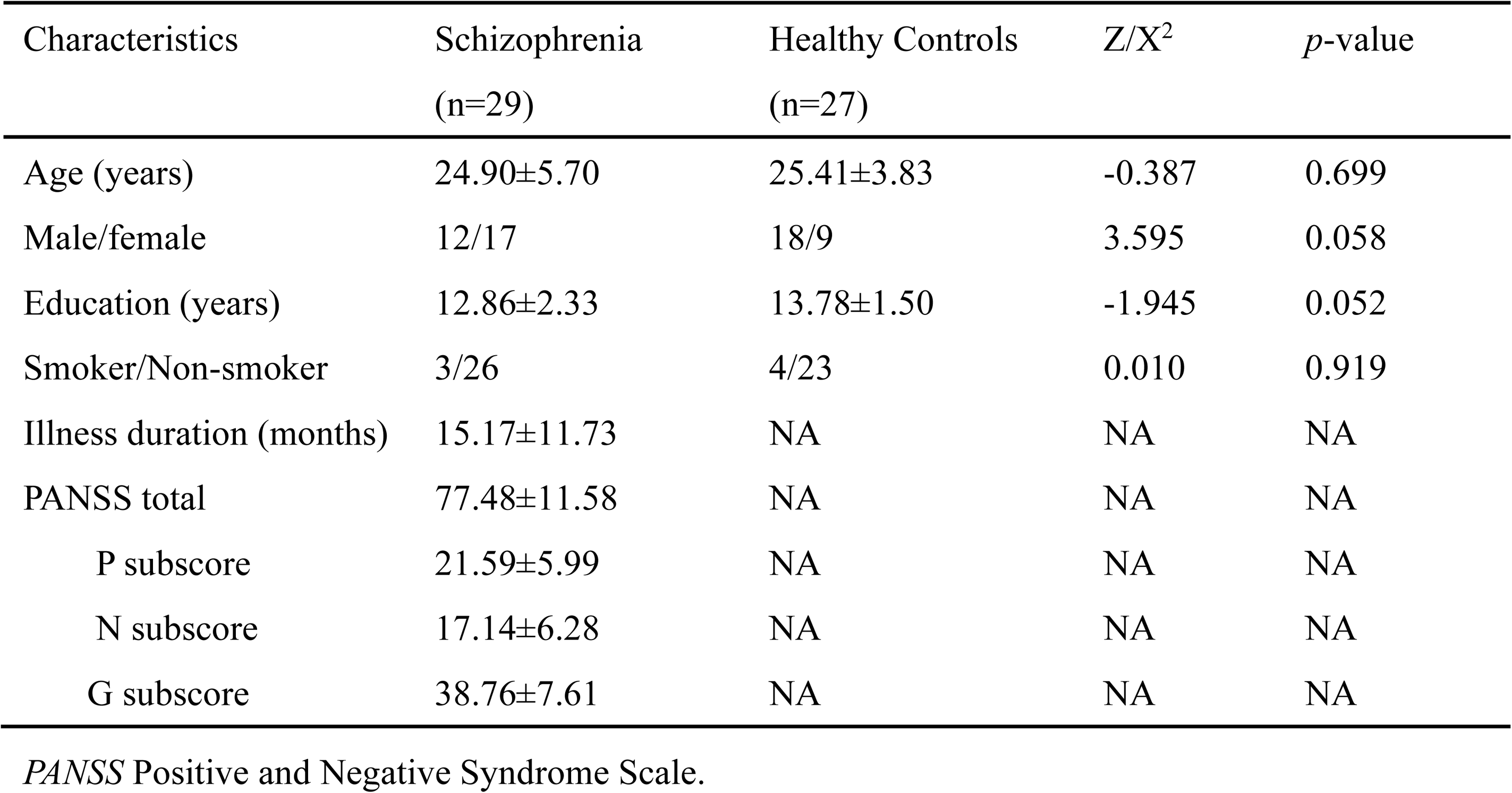
Demographic and clinical characteristics in the participants receiving monocytic subtyping by flow cytometry.

**Supplementary Table 4B.**
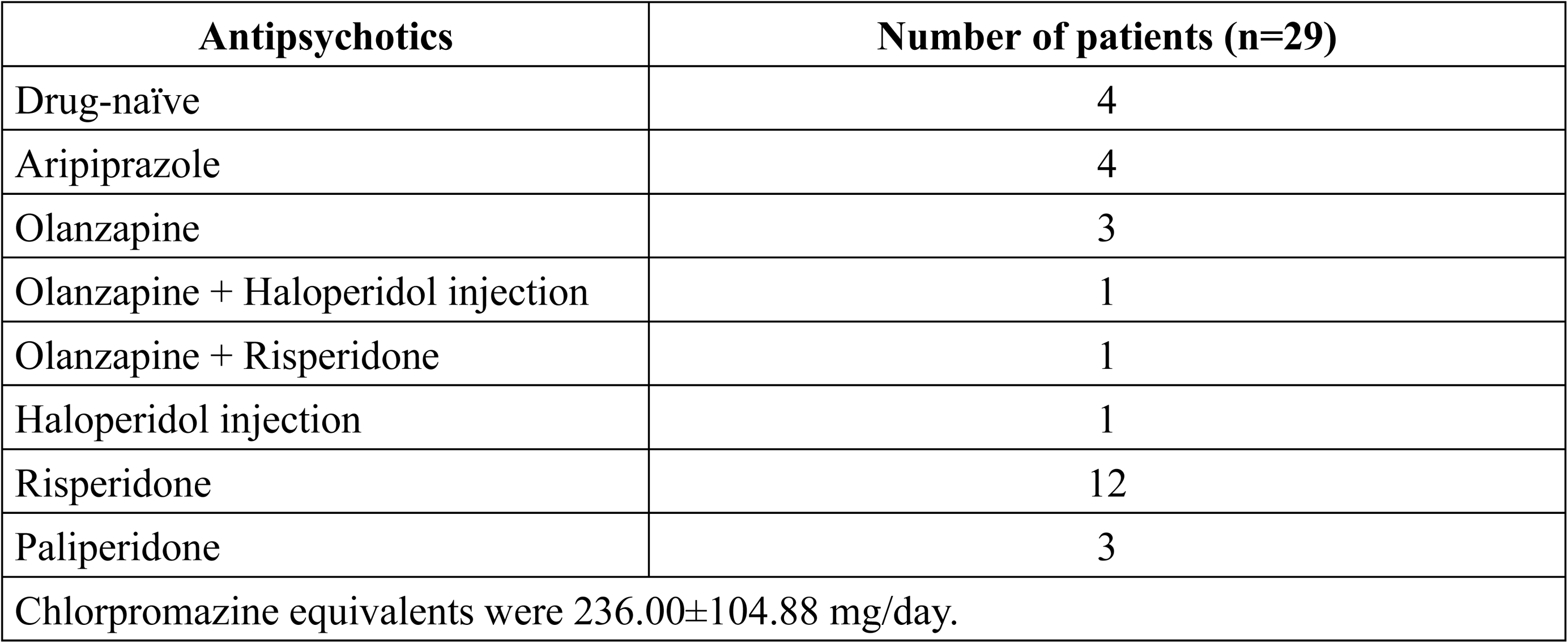
Distribution of antipsychotics at the time of enrollment in the patients receiving monocytic subtyping by flow cytometry.

**Supplementary Table 4C.**
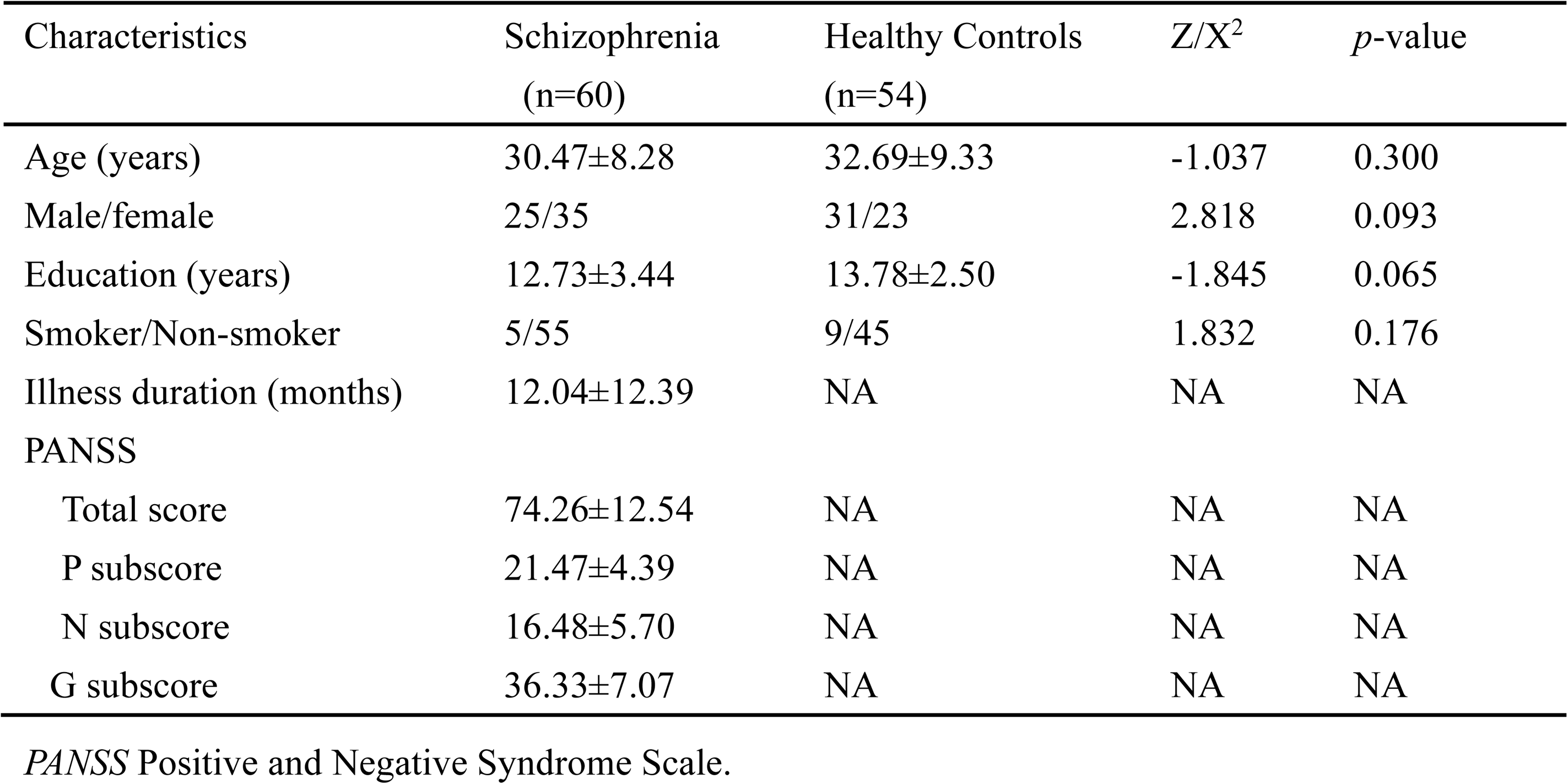
Demographic and clinical characteristics in participants undergoing brain imaging.

**Supplementary Table 4D.**
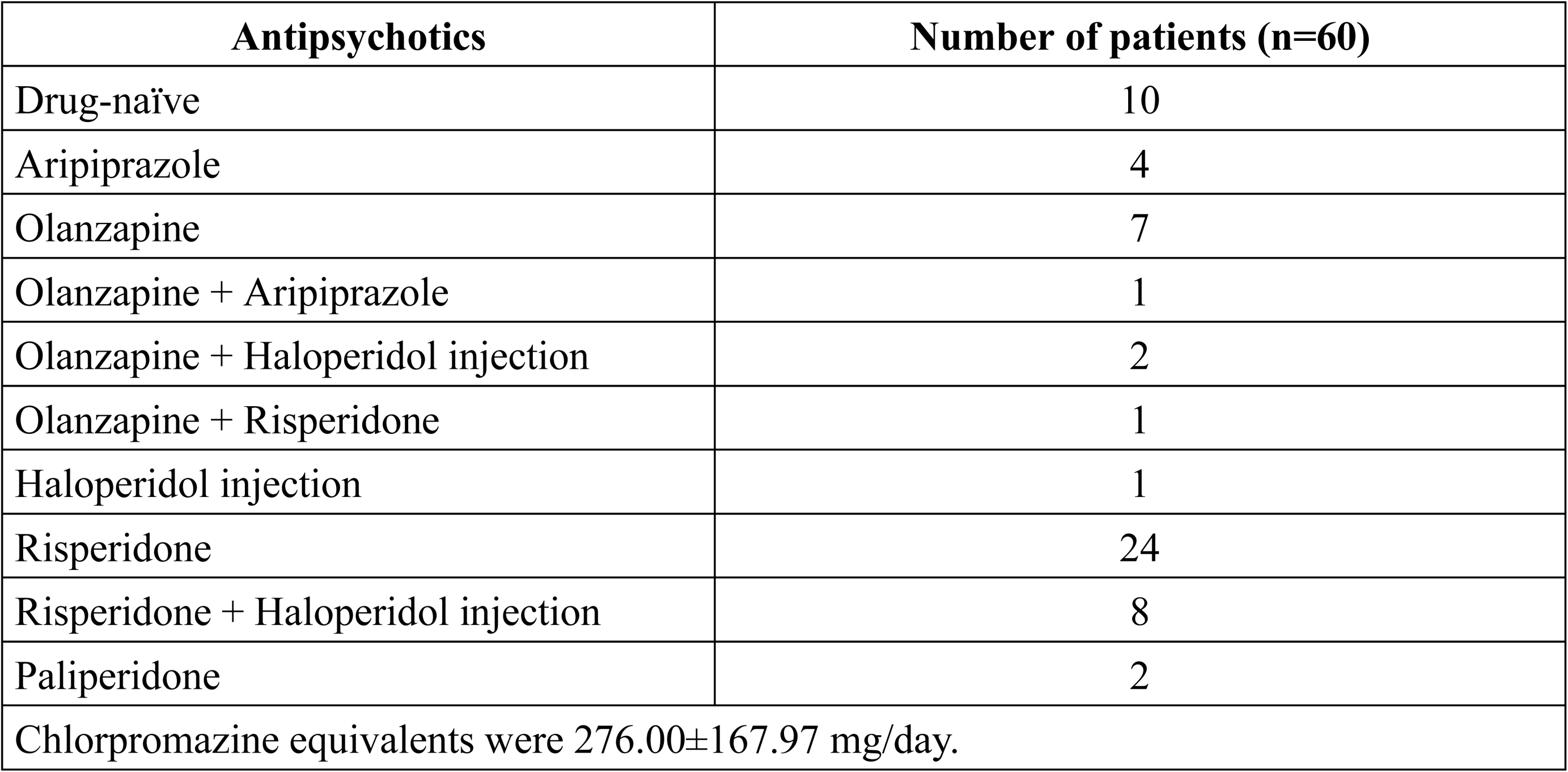
Distribution of antipsychotics at the time of enrollment in patients undergoing brain imaging.

**Supplementary Table 5.**
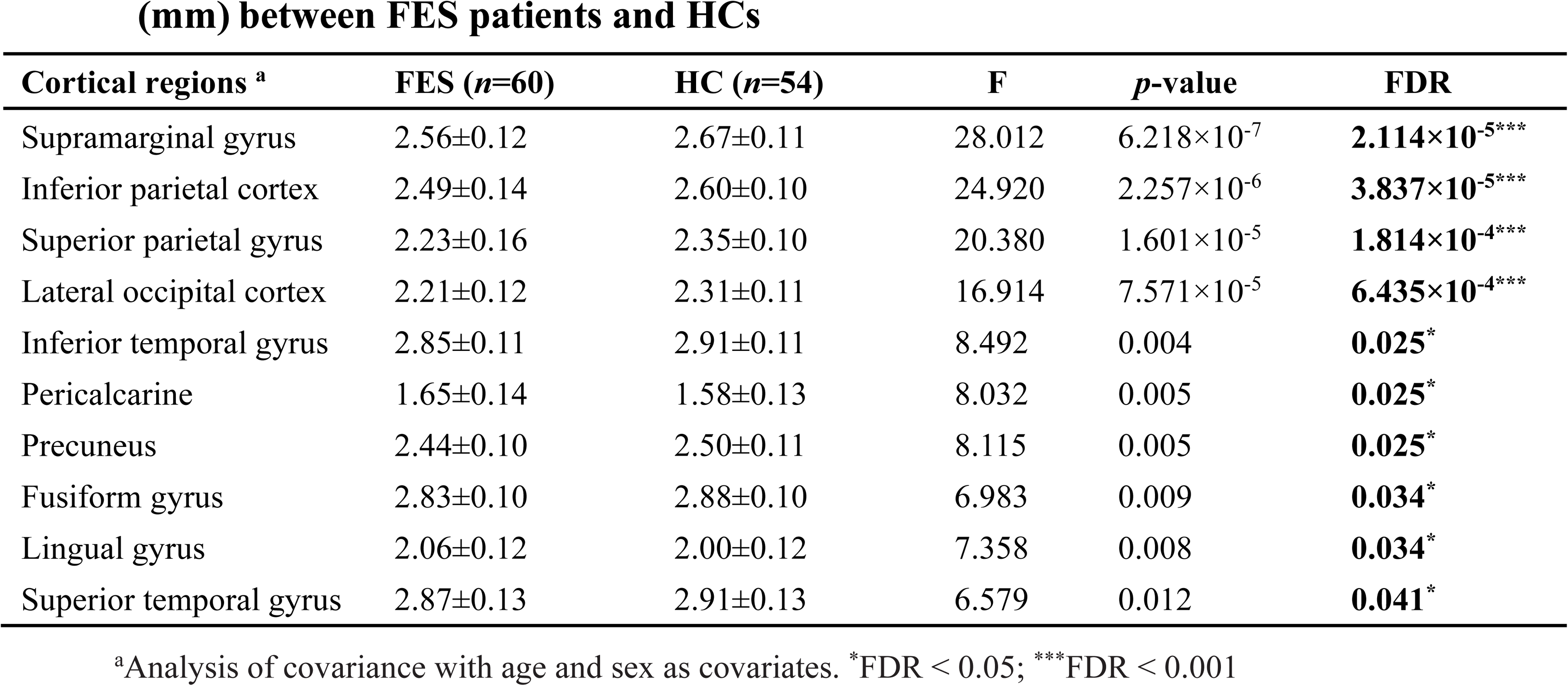
Brain regions with significant differences in cortical thickness (mm) between FES patients and HCs.

## Notes

### Competing Interest Statement

The authors have declared no competing interest.

### Funding Statement

This work was supported by the Academy of Finland research grant No. 273108, the Estonian Research Council-European Union Regional Developmental Fund Mobilitas Pluss Program No. MOBTT77, and the National Natural Science Foundation of China (81761128021 and 82001415).

### Author Declarations

This study complied with the Declaration of Helsinki with regard to an investigation in humans, and the study protocol was approved by the Medical Ethical Committee of Beijing Huilongguan Hospital. All of the participants gave written informed consent before the initiation of study procedures.

